# Transcriptomic Profiling in Skeletal Muscle Identifies Associations With Knee Osteoarthritis: the Study of Muscle, Mobility and Aging (SOMMA)

**DOI:** 10.64898/2026.04.14.26350906

**Authors:** Daniel S. Evans, Tyler A. Mansfield, Gina M. Many, Tyler J. Sagendorf, Samaneh Farsijani, Bret H. Goodpaster, Lauren M. Sparks, Nancy E. Lane

**Affiliations:** Center for Aging Science, Sutter Health Research Institute, San Francisco, CA; Department of Epidemiology and Biostatistics, University of California, San Francisco, CA; Biological Sciences Division, Pacific Northwest National Laboratory, Richland, WA; University of Pittsburgh, Pittsburgh, PA; Translational Research Institute, AdventHealth, Orlando, FL; Department of Internal Medicine, U.C. Davis Health, Sacramento, CA

## Abstract

**Objectives:** The association between skeletal muscle gene expression and knee osteoarthritis (OA) was examined among older adult participants of the Study of Muscle, Mobility and Aging (SOMMA).

**Methods:** Inclusion criteria included knee radiographs and bulk RNA sequencing (RNAseq) in *vastus lateralis* muscle, resulting in 523 participants (56% female). Radiographic knee OA was determined by Kellgren-Lawrence (KL) grades. Differential gene expression was analyzed using a control group (KL ≤ 1, n = 326) and two nested case groups: (a) KL ≥ 2 (n = 197), (b) KL ≥ 3 (n = 112).

**Results:** Compared with controls, there were 27 and 41 genes associated (FDR ≤ 0.05) with KL ≥ 2 and KL ≥ 3, respectively, and 16 genes significantly associated in both contrasts. For 15 of the 16 genes, the association magnitude was larger with more severe OA (KL ≥ 3). Genes associated in both contrasts included brain-derived neurotrophic factor (BDNF) and interferon regulatory factor-2 (IRF2). Gene sets enriched in KL ≥ 2 and KL ≥ 3 contrasts included DNA repair and branched chain amino acid (BCAA) catabolism.

**Conclusions:** Our results in older adult SOMMA participants indicate that knee OA is associated with genes and pathways expressed in skeletal muscle that are involved in pain sensitization, BCAA catabolism, muscle function preservation, calcium transport and storage, inflammation, and extracellular matrix remodeling. Additional longitudinal studies will be needed to determine how these genes could affect the progression of knee OA.

## Introduction

Knee OA is common in older individuals, affecting 263 million individuals worldwide [1]. Individuals affected with knee OA often display reductions in skeletal muscle mass, quality, and strength, yet the mechanisms behind these changes are broadly unknown [2, 3]. Currently, physical activity appears to be the only therapy for muscle loss associated with knee OA, but its effectiveness declines with advanced age, and is only moderately successful [4, 5]. It is well appreciated that tissues outside of the joint contribute to the development and progression of OA, but little is known about the molecular factors in muscle that might play a role [6].

The few studies that have explored skeletal muscle gene expression associations with knee OA have either been limited in sample size, used targeted gene expression panels, or employed experimental designs that restricted the inferences that could be drawn. Noehren et al., performed a cellular analysis and a targeted gene expression study assaying Connective Tissue Growth Factor (CTGF) and Transforming Growth Factor beta (TGFβ) from vastus lateralis biopsied from knee OA participants (n = 24) and controls (n = 15). Knee OA subjects had significantly greater extracellular matrix (ECM) content, lower density of satellite cells, and higher profibrotic gene expression (TGFβ) than controls [6]. The authors hypothesized that impaired satellite cell density, high profibrotic gene expression, and a slow-to-fast fiber type transition may contribute to reduced muscle quality in OA, but their conclusions about gene expression were limited to the targeted gene panel. More recently, Drummer et al., profiled the skeletal muscle transcriptome using RNAseq from the muscle overlaying the surgical joint and the contralateral or control limb in 20 females undergoing either total hip or knee replacement and identified differentially expressed genes related to inflammation and lipid metabolism [7]. While these results point to the importance of muscle gene expression in OA, many questions remain. The study design of Drummer et al. did not include radiographic knee OA presence or severity in the contralateral control joint, so the genes identified might not represent a distinct contrast between joints with and without OA. Moreover, Drummer et al. examined end-stage OA in females who underwent joint replacement, and thus does not address gene expression associations with mild to moderate knee OA, nor does it include males. To date, a large-scale genome-wide transcriptomic analysis of skeletal muscle and knee OA in males and females has not been conducted.

In the present study, we performed an analysis of RNAseq transcriptomic data obtained from a biopsy of the vastus lateralis muscle in 523 male and female participants that also had knee radiographs from the Study of Muscle, Mobility and Aging (SOMMA). We characterized gene expression between participants without radiographic knee OA and those with moderate to severe radiographic knee OA to test the hypothesis that genes and regulatory pathways expressed in skeletal muscle are associated with radiographic knee OA in males and females with and without knee pain.

## Methods

### Study design

SOMMA aims to determine the biologic processes that contribute to changes in mobility and fitness with aging [8]. There are two clinical sites (University of Pittsburgh and Wake Forest University School of Medicine), a biorepository (AdventHealth Translational Research Institute), and a coordinating center (San Francisco Coordinating Center, California Pacific Medical Center Research Institute). Study participants were included in SOMMA if they were ≥70 years old and able to complete a 400 m walk at ≥0.6 m/s. Study participants had to be free of life-threatening disease and had no contraindications to magnetic resonance imaging (MRI) or tissue collection. Study exclusion criteria include BMI ≥ 40 kg/m^2^, or if participants reported an inability to walk one-fourth of a mile or climb a flight of stairs, an active malignancy, or advanced chronic disease [8].

This ancillary cross-sectional study enrolled SOMMA participants who returned for the first annual visit and consented to completing a standing knee radiograph. Participants provided written informed consent, and the study was approved by the Western Institutional Review Board Copernicus Group (no. 20180764) for all participating sites [8, 9]. Data from the November 2024 SOMMA release was analyzed.

### Radiographic and symptomatic OA

Bilateral, fixed-flexion, weight-bearing posteroanterior knee radiographs were obtained at the SOMMA one-year follow-up visit. The presence and severity of knee OA was determined with Kellgren-Lawrence (KL) grades (scale 0–4), and the extent of medial and lateral joint space narrowing were scored centrally based on the Osteoarthritis Research Society International atlas of individual radiographic features. Reproducibility of the KL grade readings had a weighted kappa for interrater reliability of 0.75 (95% confidence interval [CI] 0.69–0.79] [9].

Based on tibiofemoral KL grades of the same knee from which the muscle biopsy was obtained, participants were assigned to three groups: KL ≤ 1 (normal to doubtful, controls), KL ≥ 2 (mild to severe), and KL ≥ 3 (moderate to severe). Self-reported knee pain was defined as knee pain occurring at least monthly in the same knee from which the muscle biopsy was obtained [9].

### General study measurements

SOMMA baseline assessments included self-reported information on birth date, sex, race, ethnicity, education, substance use, medical history, and prescription medications [8]. Gait speed was assessed with a 400-meter walk. Daily step count was measured using an accelerometer (ActiGraph GT9X) with data processed using the GGIR R package (version 3.1.1) [10]. Height was measured on stadiometers, and weight was measured using a digital scale. Total abdominal adipose tissue mass was quantified from whole-body MRI as the summed abdominal adipose tissue volume within the abdominal compartment (subcutaneous + visceral) using automated segmentation algorithms from AMRA Researcher® (AMRA Medical AB, Linköping Sweden). Anterior thigh fat-free muscle volume (same leg as muscle biopsy, anterior compartment) was also derived from MRI by AMRA Researcher® using automated segmentation to identify voxels with low fat fraction and summing their volume to give fat-free (viable) muscle volume [8].

### Muscle biopsy collection, RNA isolation, and sequencing

Skeletal muscle biopsy collection and processing, RNA isolation, library preparation, and sequencing was performed at the baseline SOMMA visit as previously described [11, 12]. To process the sequence data, we first removed adaptors of the sequenced reads using the cutadapt software (v3.4). The cleaned reads were then aligned to the Genome Reference Consortium Human Build 38 (GRCh38) Ensembl release 112 using the STAR software (v2.7.11b) [13]. Genome-aligned reads were remapped to the transcriptome using STAR and the Ensembl v112 gtf. Duplicated transcriptome-aligned reads were further marked and removed using the Picardtools software (v3.1.1). Expression count data were obtained using the RSEM software (v1.3.3) [14]. RSEM transcript quantifications were then summarized to the gene level with tximport [15].

### Statistical Analysis

Baseline demographic, clinical, and physical performance characteristics were summarized for the full analytic sample and stratified by KL grade of the same knee from which the muscle biopsy was obtained (KL ≤ 1, KL ≥ 2, and KL ≥ 3). Group differences (KL ≤ 1 vs KL ≥ 2 and KL ≤ 1 vs KL ≥ 3) for continuous variables were evaluated using t-tests for normally distributed variables and Wilcoxon rank-sum tests for skewed variables (age and daily step count). Group differences for categorical variables were evaluated using chi-squared tests. All comparisons were also conducted separately for men and women.

Gene expression associations with KL grades were analyzed using DESeq2 (version 1.46.0) [16]. Models were adjusted for age, sex, clinic site, race/ethnicity (non-Hispanic White vs other), weight, waist circumference, total abdominal adiposity, anterior thigh fat-free muscle volume (from the same knee as the biopsy), daily step count, nonsteroidal anti-inflammatory drug (NSAID) use (oral and topical NSAID preparations taken within 30 days before the clinic visit), and sequencing batch. DESeq2 fits a negative binomial generalized linear model to estimate differential expression and uses Wald tests to assess the significance of regression coefficients. Genes with fewer than 10 samples contributing at least 10 total counts were excluded prior to modeling. Multiple testing correction was applied using the Benjamini–Hochberg false discovery rate (FDR), with statistical significance defined as FDR < 0.05. Two primary contrasts were examined (KL ≤ 1 vs KL ≥ 2 and KL ≤ 1 vs KL ≥ 3) where a positive log2 fold change indicates higher expression in the case group (KL ≥ 2 or KL ≥ 3).

As a sensitivity analysis, genes that were significantly associated in both primary contrasts were further examined in sex-stratified models. Additionally, two secondary case definitions were analyzed in sensitivity analyses: KL ≥ 2 with knee pain and KL ≥ 3 with knee pain.

Pathway enrichment analysis was performed using the Pre-Ranked Correlation Adjusted MEan RAnk gene set test (CAMERA-PR). Gene sets were obtained from the Reactome Canonical Pathways and Gene Ontology subcollections of the Molecular Signatures Database (MSigDB), as well as human gene sets from the Ma’ayan Laboratory’s CellMarker_2024.txt (https://maayanlab.cloud/Enrichr/#libraries). The analysis was implemented using the camera function in limma, which applies a modified two-sample t-test that accounts for inter-gene correlations. In all analyses, we used the default inter-gene correlation parameter of 0.01 provided by limma. Wald z-statistics from the DESeq2 differential expression analysis were provided as the ranking metric. Multiple testing correction was performed using the Benjamini-Hochberg procedure separately for each contrast, controlling the false discovery rate across all tested gene sets.

## Results

There were 523 SOMMA participants (56% female) with bulk RNAseq from the biopsy of the vastus lateralis, knee radiographs, and a complete set of covariates (Figure 1). Knee OA controls (n = 326) were defined as KL grade ≤ 1 (Table 1). The controls were compared with two knee OA case groups: KL grade ≥ 2 (n = 197) and the more severe KL grade ≥ 3 (n = 112) (Table 1). The median age of the study participants was 75.3 years (Q1, Q3 = 72.7, 79.1) and did not differ across knee OA groups. Weight, BMI, and waist circumference were higher in both knee OA case groups compared with controls, as was total adipose tissue volume (Table 1). Fat-free anterior thigh muscle volume from the same leg as the biopsy was not significantly different between knee OA groups, but the means were lower in both knee OA case groups compared with knee OA controls (Table 1). Consistent with expectations for knee OA, walking speed and daily step count were lower in both knee OA case groups compared with controls, and monthly reported knee pain was more frequent in both knee OA case groups compared with controls (Table 1). These observations were largely consistent in sex-stratified analysis (Table S1).

**Figure 1.**
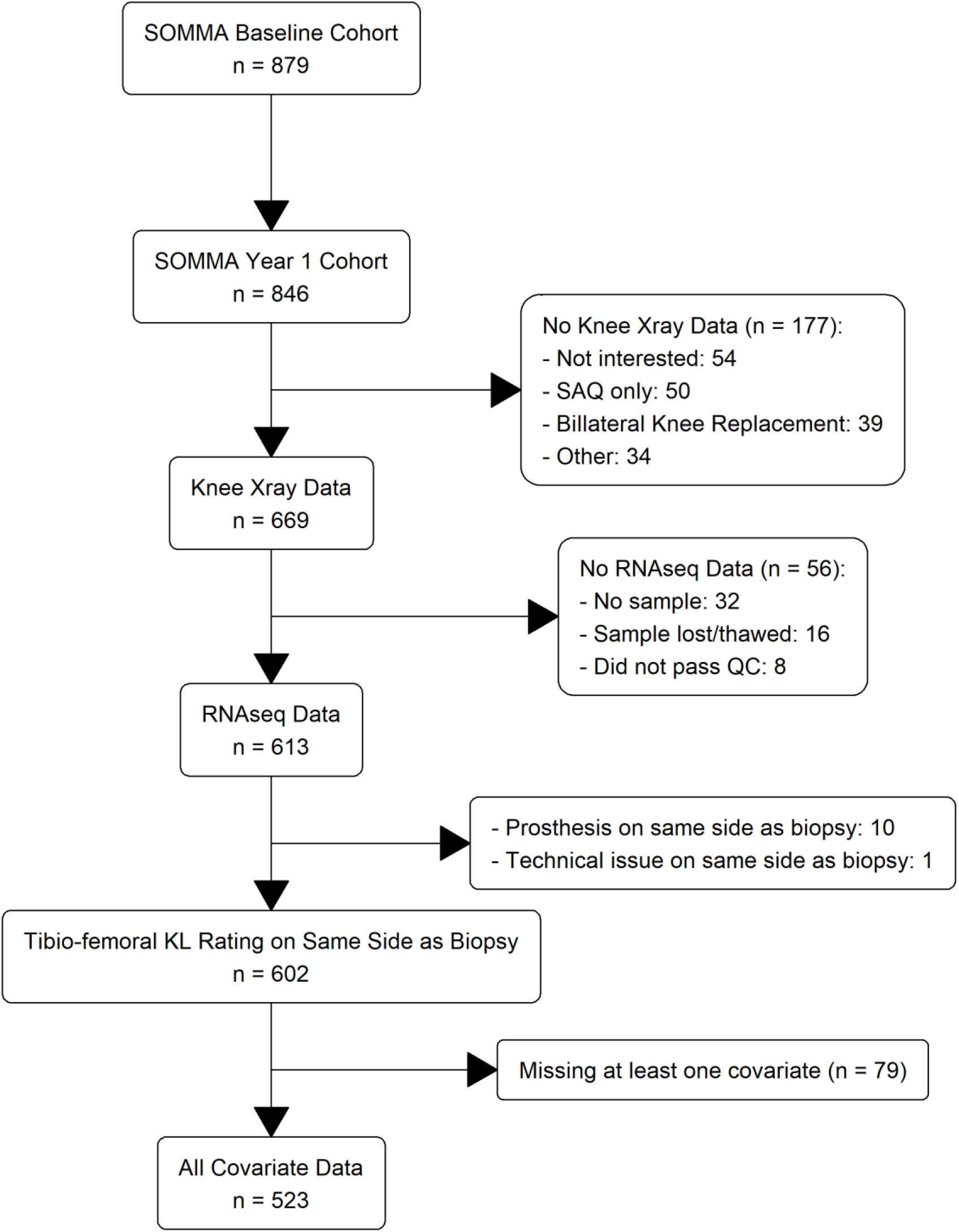
Analytic Cohort Consort Diagram.

**Table 1.** Study Participant characteristics.

| Variable | OVERALL<br>N=523 | KL ≤ 1<br>N=326 | KL ≥ 2<br>N=197 | KL ≥ 3<br>N=112 |
| --- | --- | --- | --- | --- |
| Age, years | 75.3<br>(72.7, 79.1) | 75.1<br>(72.7, 78.5) | 75.6<br>(72.9, 80.4) | 75.4<br>(72.1, 80.3) |
| Female, n (%) | 292 (55.8) | 174 (53.4) | 118 (59.9) | 70 (62.5) |
| Site, Pittsburgh | 227 (43.4) | 128 (39.3) | 99 (50.3)* | 59 (52.7)* |
| Non-Hispanic White | 456 (87.2) | 289 (88.7) | 167 (84.8) | 93 (83.0) |
| Height, m | 1.66 ± 0.10 | 1.66 ± 0.10 | 1.66 ± 0.10 | 1.65 ± 0.10 |
| Weight, kg | 75.4 ± 14.8 | 73.9 ± 14.6 | 77.8 ± 15.0** | 77.5 ± 14.7* |
| BMI, kg/m<sup>2</sup> | 27.3 ± 4.3 | 26.6 ± 4.2 | 28.3 ± 4.3*** | 28.3 ± 4.2*** |
| Waist circumference, cm | 94.2 ± 13.1 | 92.8 ± 13.4 | 96.4 ± 12.4** | 96.0 ± 11.9* |
| Total Adipose Tissue Volume (VAT+SAT), L | 11.59 ± 4.33 | 11.12 ± 4.35 | 12.36 ± 4.21** | 12.27 ± 3.76* |
| Anterior Thigh Fat-Free Muscle Volume, L † | 1.54 ± 0.44 | 1.55 ± 0.44 | 1.51 ± 0.44 | 1.47 ± 0.44 |
| Walking Speed for 400m walk, m/s | 1.07 ± 0.17 | 1.10 ± 0.16 | 1.02 ± 0.18*** | 1.01 ± 0.18*** |
| Daily Step Count | 4937<br>(3071, 7071) | 5228<br>(3448, 7177) | 4168<br>(2751, 7063)* | 4183<br>(2693, 6581)* |
| NSAID Use | 41 (7.8) | 21 (6.4) | 20 (10.2) | 15 (13.4)* |
| Knee Pain† | 177 (34.0) | 76 (23.5) | 101 (51.3)*** | 65 (58.0)*** |

\*= P<0.05, \*\* = p<0.005 , \*\*\* = p<0.0005
† Same side as muscle biopsy for RNAseq

### Single gene results

Compared with controls, there were 27 and 41 differentially expressed genes (FDR ≤ 0.05) in the KL ≥ 2 and KL ≥ 3 contrasts with controls, respectively (Figure 2 and Table S2). Sixteen of these genes were significantly associated in both contrasts, and the effect directions were the same for all 16 genes (Table 2). The gene expression effect size magnitude was larger for 15 of the 16 genes in contrasts with more severe OA cases (KL ≥ 3) compared with less severe cases (KL ≥ 2), demonstrating a relationship between stronger gene expression associations with the severity of joint structural damage (Table 2).

**Figure 2.**
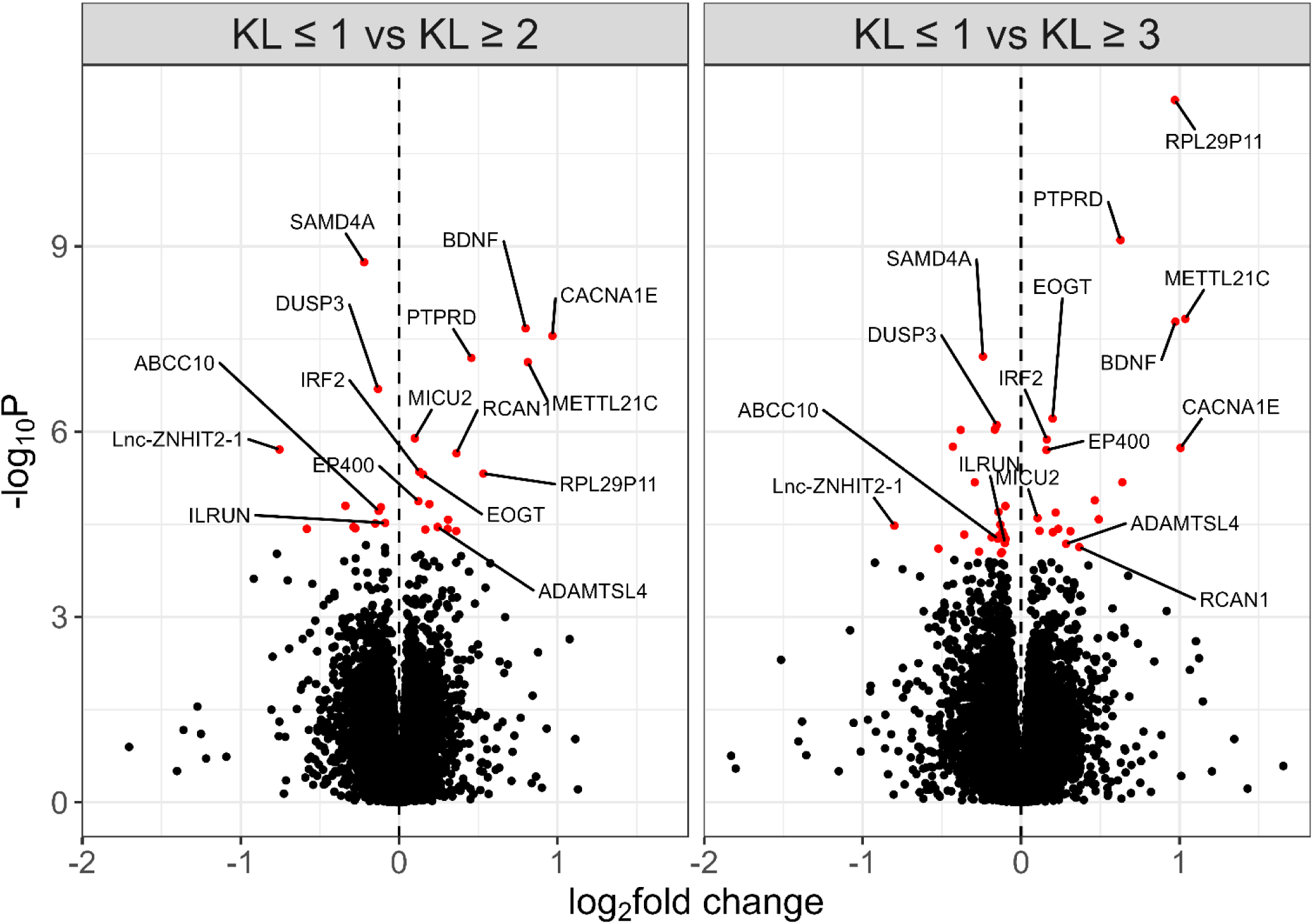
Volcano plot demonstrating the genes differentially expressed in the KL ≥ 2 group and the KL ≥ 3 group compared to KL ≤ 1. Results are expressed in log fold changes. Red points indicate genes that were statistically significant after false discovery rate correction (FDR ≤ 0.05). Annotated genes represent those that were significant (FDR ≤ 0.05) in both contrasts.

**Table 2.**
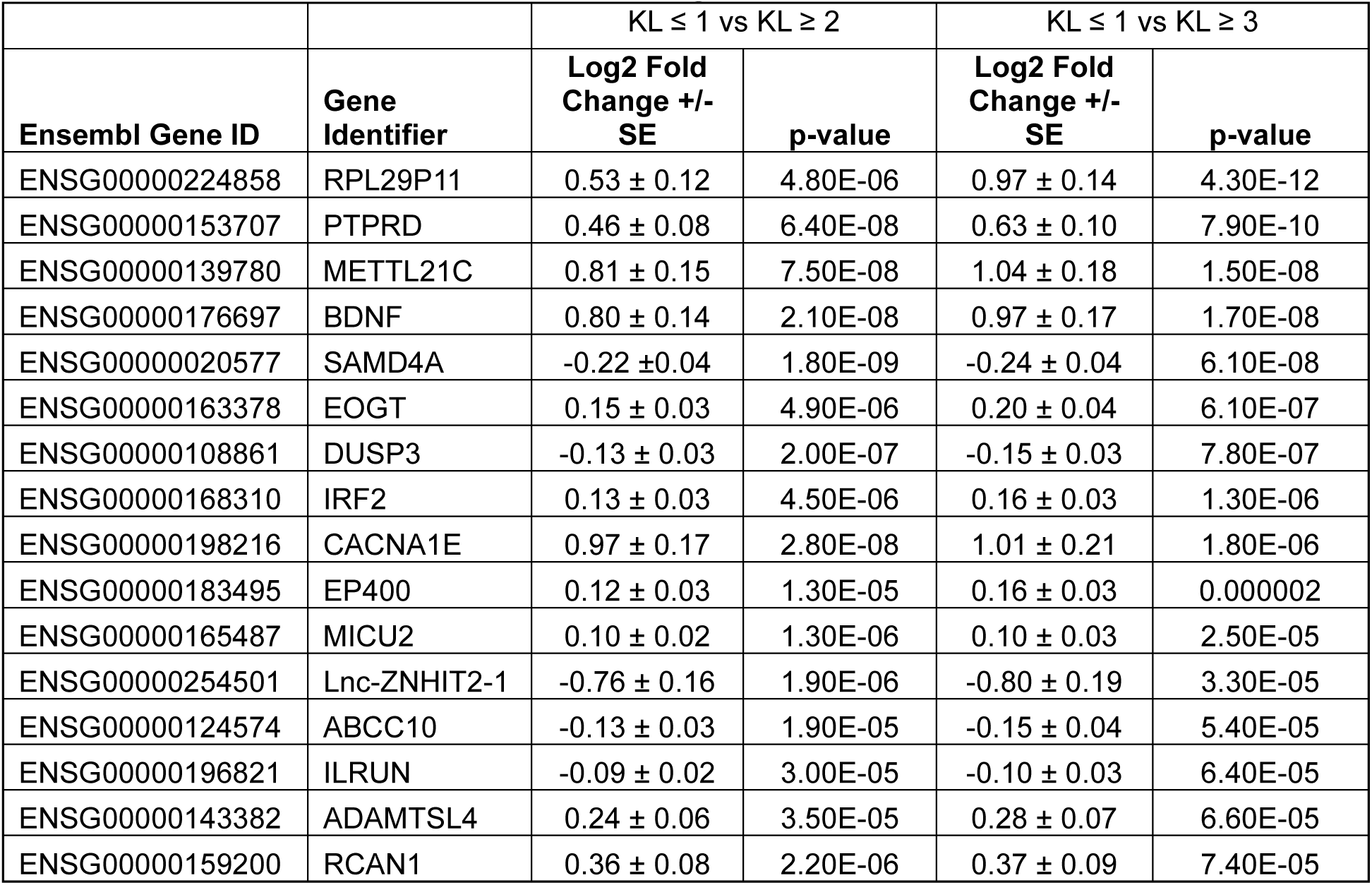
Gene expression associations significant in both contrasts.

| Ensembl Gene ID | Gene Identifier | KL $\leq 1$ vs KL $\geq 2$ | | KL $\leq 1$ vs KL $\geq 3$ | |
| --- | --- | --- | --- | --- | --- |
|  |  | Log2 Fold Change +/- SE | p-value | Log2 Fold Change +/- SE | p-value |
| ENSG00000224858 | RPL29P11 | 0.53 ± 0.12 | 4.80E-06 | 0.97 ± 0.14 | 4.30E-12 |
| ENSG00000153707 | PTPRD | 0.46 ± 0.08 | 6.40E-08 | 0.63 ± 0.10 | 7.90E-10 |
| ENSG00000139780 | METTL21C | 0.81 ± 0.15 | 7.50E-08 | 1.04 ± 0.18 | 1.50E-08 |
| ENSG00000176697 | BDNF | 0.80 ± 0.14 | 2.10E-08 | 0.97 ± 0.17 | 1.70E-08 |
| ENSG00000020577 | SAMD4A | -0.22 ± 0.04 | 1.80E-09 | -0.24 ± 0.04 | 6.10E-08 |
| ENSG00000163378 | EOGT | 0.15 ± 0.03 | 4.90E-06 | 0.20 ± 0.04 | 6.10E-07 |
| ENSG00000108861 | DUSP3 | -0.13 ± 0.03 | 2.00E-07 | -0.15 ± 0.03 | 7.80E-07 |
| ENSG00000168310 | IRF2 | 0.13 ± 0.03 | 4.50E-06 | 0.16 ± 0.03 | 1.30E-06 |
| ENSG00000198216 | CACNA1E | 0.97 ± 0.17 | 2.80E-08 | 1.01 ± 0.21 | 1.80E-06 |
| ENSG00000183495 | EP400 | 0.12 ± 0.03 | 1.30E-05 | 0.16 ± 0.03 | 0.000002 |
| ENSG00000165487 | MICU2 | 0.10 ± 0.02 | 1.30E-06 | 0.10 ± 0.03 | 2.50E-05 |
| ENSG00000254501 | Lnc-ZNHIT2-1 | -0.76 ± 0.16 | 1.90E-06 | -0.80 ± 0.19 | 3.30E-05 |
| ENSG00000124574 | ABCC10 | -0.13 ± 0.03 | 1.90E-05 | -0.15 ± 0.04 | 5.40E-05 |
| ENSG00000196821 | ILRUN | -0.09 ± 0.02 | 3.00E-05 | -0.10 ± 0.03 | 6.40E-05 |
| ENSG00000143382 | ADAMTSL4 | 0.24 ± 0.06 | 3.50E-05 | 0.28 ± 0.07 | 6.60E-05 |
| ENSG00000159200 | RCAN1 | 0.36 ± 0.08 | 2.20E-06 | 0.37 ± 0.09 | 7.40E-05 |

Sex-stratified analysis was performed to examine whether gene expression associations with knee OA might differ by sex. Sex-stratified results (Table S3) were largely consistent with sex-adjusted results (Table 2). Two genes that were significantly associated in sex-adjusted results for both contrasts (RPL29P11 and RCAN1) were not nominally significant (P ≥ 0.05) in men, suggesting that the associations could be sex-specific (Table S3). However, the 95% confidence intervals (CI) of the sex-specific gene expression associations with knee OA overlapped, indicating that the effect estimates did not significantly differ by sex. For RPL29P11, the 95% CI overlapped for contrast 1 (controls vs KL ≥ 2) in females (0.53, 1.16) and males (-0.72, 0.76) and for contrast 2 (controls vs KL ≥ 3) in females (0.98, 1.72) and males (-0.95, 0.98). RPL29P11 is the third lowest expressed gene of the 16 differentially-expressed genes across both contrasts, which could account for sex-specific variability in effect estimates. RCAN1 also showed apparently different knee OA associations by sex, and was also highly significantly associated with knee OA in females but not males (Table S3). As was the case with RPL29P11, the 95% CI of RCAN1’s sex-specific knee OA associations overlapped for both contrasts. RCAN1’s 95% CI overlapped for contrast 1 (controls vs KL ≥ 2) in females (0.26, 0.67) and males (-0.01, 0.44) and for contrast 2 (controls vs KL ≥ 3) in females (0.22, 0.70) and males (-0.04, 0.55).

To examine whether self-reported knee pain impacted the gene expression associations, we added knee pain to the two case definitions, such that we created two new case groups: KL ≥ 2 and pain, and KL ≥ 3 and pain. No new gene associations were identified when adding pain to the case definitions (Figure 3). In fact, there were fewer gene expression associations with case definitions that included pain, which could be due, at least in part, to the smaller sample size of case definitions with pain.

**Figure 3.**
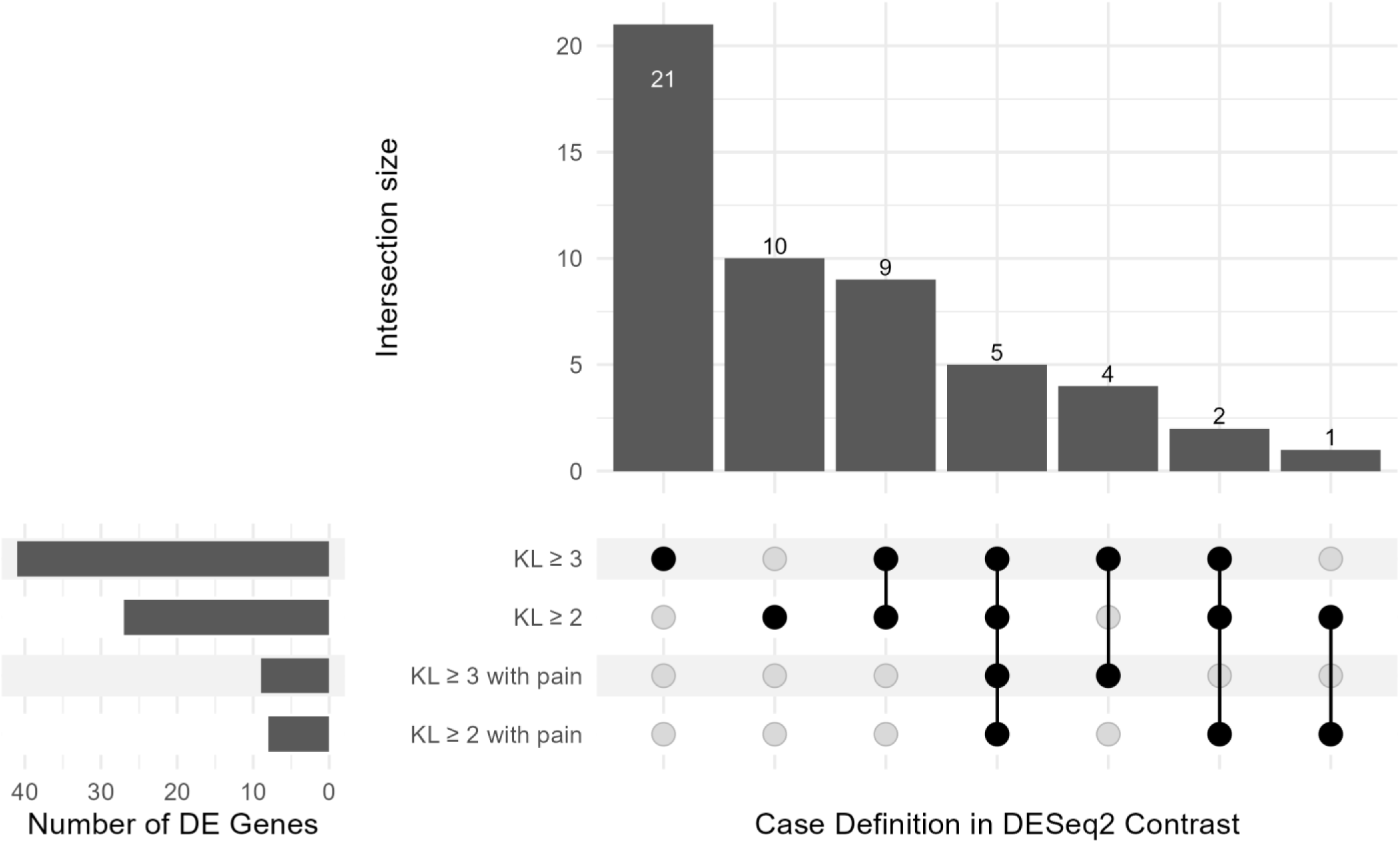
Upset plot showing the intersection of significantly associated genes across different contrasts. All groups are compared to KL ≤ 1. Numbers on each bar represents the number of genes in each intersection.

Brain-derived neurotrophic factor (BDNF), one of the significantly upregulated genes in both contrasts (Table 2), has been previously reported to be associated with OA-related pain in clinical studies and laboratory models [17-20]. To examine whether our results were consistent with that finding, we examined whether BDNF’s association became stronger when pain was included in our OA case definition. Indeed, BDNF’s association with KL ≥ 2 with pain (log_2_FC = 1.01, P-value = 9.6x10^-9^) was stronger than its association with KL ≥ 2 without considering pain (log_2_FC = 0.80, P-value = 2.1x10^-8^). BDNF’s association was also stronger with pain when using the more severe OA definition (KL ≥ 3 with pain: log_2_FC = 1.13, P-value = 7x10^-8^; KL ≥ 3 without considering pain: log_2_FC = 0.97, P-value = 1.7x10^-8^). While BDNF’s association effect size with knee OA was larger with pain, the 95% confidence intervals overlapped, so the pain-specific associations were not significantly different from knee OA without considering pain.

### Pathway enrichment

There were 20 gene sets significantly enriched (FDR ≤ 0.05) in KL ≥ 2 and KL ≥ 3 contrasts, which included pathways related to DNA replication and repair, fatty acid metabolism, branched chain amino acid (BCAA) catabolism, and chondrocyte cell identity (Table 3). The test statistics were largely similar in both contrasts, with the exception of the two downregulated gene sets, in which the enrichment test-statistics were nearly double in magnitude in the KL ≥ 3 contrast compared with the KL ≥ 2 contrast.

**Table 3.** Gene-set enrichment significant in both contrasts.

| Gene set | KL ≥ 2 contrast | | KL ≥ 3 contrast | |
| --- | --- | --- | --- | --- |
|  | t-statistic | P-value | t-statistic | P-value |
| GOMF NADPLUS BINDING | 5.26 | 1.47E-07 | 3.82 | 0.000136 |
| GOBP FATTY ACID BETA OXIDATION USING ACYL-COA DEHYDROGENASE | 4.86 | 1.17E-06 | 3.51 | 0.000443 |
| GOBP BRANCHED CHAIN AMINO ACID CATABOLIC PROCESS | 4.45 | 8.61E-06 | 3.56 | 0.000377 |
| GOBP CELL CYCLE DNA REPLICATION | 4.43 | 9.57E-06 | 3.88 | 0.000104 |
| GOBP LIPID OXIDATION | 4.42 | 1E-05 | 3.49 | 0.000481 |
| GOBP DNA CONFORMATION CHANGE | 4.39 | 1.16E-05 | 3.52 | 0.000431 |
| REACTOME BRANCHED CHAIN AMINO ACID CATABOLISM | 4.37 | 1.24E-05 | 3.58 | 0.000344 |
| GOMF DNA REPLICATION ORIGIN BINDING | 4.36 | 1.3E-05 | 4.56 | 5.04E-06 |
| REACTOME EUKARYOTIC TRANSLATION ELONGATION | -4.25 | 2.17E-05 | -8.50 | 2E-17 |
| CELLMARKER Homeostatic Chondrocyte Articular Cartilage Human | 4.24 | 2.23E-05 | 4.23 | 2.32E-05 |
| GOCC ENDOPLASMIC RETICULUM EXIT SITE | 4.19 | 2.86E-05 | 4.03 | 5.59E-05 |
| GOMF OXIDOREDUCTASE ACTIVITY ACTING ON THE CH-CH GROUP OF DONORS WITH A FLAVIN AS ACCEPTOR | 4.17 | 3.11E-05 | 3.51 | 0.000443 |
| GOMF DNA HELICASE ACTIVITY | 4.10 | 4.16E-05 | 4.00 | 6.44E-05 |
| GOMF ATP DEPENDENT ACTIVITY ACTING ON DNA | 4.06 | 5E-05 | 4.13 | 3.67E-05 |
| GOBP REGULATION OF DNA TEMPLATED DNA REPLICATION | 4.02 | 5.9E-05 | 3.58 | 0.000339 |
| REACTOME RESPONSE OF EIF2AK4 GCN2 TO AMINO ACID DEFICIENCY | -4.01 | 6.04E-05 | -7.99 | 1.37E-15 |
| GOBP SHORT CHAIN FATTY ACID METABOLIC PROCESS | 3.99 | 6.57E-05 | 3.54 | 0.000395 |
| REACTOME ACTIVATION OF THE PRE-REPLICATIVE COMPLEX | 3.96 | 7.61E-05 | 3.62 | 0.000291 |
| GOBP RECOMBINATIONAL REPAIR | 3.81 | 0.00014 | 3.73 | 0.000192 |
| GOMF SINGLE STRANDED DNA BINDING | 3.76 | 0.000169 | 3.41 | 0.000642 |

## Discussion

In our analysis of older adult SOMMA participants, we found multiple differences in skeletal muscle gene expression in study participants with knee OA compared to those without knee OA. The functions of the genes and pathways associated with knee OA in both contrasts included pain sensitization, BCAA catabolism, muscle function preservation, calcium transport and storage, inflammation, and extracellular matrix remodeling. For 15 of the 16 genes associated with both contrasts, the association effect sizes were larger for more severe knee OA, demonstrating a strong relationship between expression association strength and severity of joint structural damage. There were few sex differences, and most results did not greatly differ with knee pain, with the exception of BDNF. Moderate to severe knee OA (KL ≥ 2) was associated with significantly increased expression of several genes, including RPL29P11, METTL21C, BDNF, PTPRD, IRF2, CACNA1E, EOGT, and ADAMTSL4, while moderate to severe knee OA was associated with significantly reduced expression of SAMD4A, DUSP3, and Lnc-ZNHIT2-1. Research indicates links between some of the listed genes and skeletal muscle function, particularly BDNF, SAMD4A, DUSP3, PTPRD, CACNA1E, and ADAMTSL4, with many findings derived from a combination of human and laboratory animal studies. We also report that expression of RPL29P11 and Lnc-ZNHIT2-1 are associated with knee OA, but little is known regarding their function in skeletal muscle or OA. Our results highlight the importance of the molecular landscape of muscle to understand the development of OA. Moreover, genes and pathways we identified can suggest candidate targets for future therapeutic approaches for knee OA.

The association between BDNF expression and OA provides an insight into the relationship between skeletal muscle and pain. BDNF is a myokine that enhances muscle health by promoting fiber-type switching towards endurance (slow-twitch) fibers, improving mitochondrial function, aiding muscle repair (satellite cell differentiation), boosting fat burning (lipid oxidation), and strengthening neuromuscular junctions (NMJs); thus, playing a key role in metabolic flexibility and muscle plasticity [21, 22]. In addition to BDNF’s beneficial effects on muscle, BDNF can also enhance pain sensitization through its actions in the central and peripheral nervous system [23]. Pain in OA-affected joints has been shown to be strongly related to BDNF in clinical and experimental studies [17-20]. Mechanistically, rodent models indicate that BDNF binding to its receptor trkB in nerves innervating the knee joint can induce pain sensitization [17]. These previous studies of OA-related pain have focused on BDNF levels in serum and synovial fluid. Our results are the first to demonstrate that BDNF expressed in muscle is also associated with OA and OA-related pain. A number of proinflammatory cytokines and adipokines released from adipose tissue have been discovered to contribute to OA-related pain [24], and our results provide early clues about molecular factors derived from skeletal muscle related to OA and OA-related pain.

Pathway enrichment results provide some clues about the metabolic states that could impact BDNF expression. We found that branched chain amino acid (BCAA) catabolism was positively enriched among OA cases. Moreover, a pathway representing the GCN2-mediated response to amino acid deficiency was downregulated, further supporting the role of amino acid levels and metabolism in muscle for the development of OA. Exercise can induce BCAA catabolism in skeletal muscle, which in turn can activate the mTORC1 pathway [25, 26]. BCAAs can also promote BDNF expression, which can be mediated by mTORC1 signaling [26]. Thus, our results support the notion that muscle in individuals with knee OA appear to utilize alternative energy sources, like BCAAs, which can activate mTORC1 signaling resulting in higher BDNF expression. BDNF expression in muscle can be beneficial for muscle in a stressed state, but it can also have negative consequences by increasing joint pain.

In addition to BDNF, we also report that higher levels of Protein Tyrosine Phosphatase Receptor Type D (PTPRD) are associated with higher odds of knee OA. PTPRD has been reported to influence pain related to OA and other causes, providing additional support for the role of muscle-expressed molecular factors contributing to OA-related pain [17, 27-29]. PTPRD has been shown to affect body weight and metabolism by acting as a receptor for the hormone asprosin in the brain, but its direct function within skeletal muscle tissue is not well-documented [29, 30].

Two OA-associated genes, ADAMTS like 4 (ADAMTSL4) and EGF domain specific O-linked N-acetylglucosamine transferase (EOGT), are involved in extracellular matrix (ECM) remodeling and production of cartilage components. ADAMTS-like proteins, including ADAMTSL4, are secreted proteins that regulate the extracellular matrix (ECM) of connective tissue [31]. EOGT transfers N-acetylglucosamine to extracellular proteins that mediates cell-cell and cell-ECM interactions [32].

We also identified three inflammation-related genes associated with knee OA: methyltransferase 21C, AARS1 lysine (METTL21C), inflammation and lipid regulator with UBA-like and NBR1-like domains (ILRUN), and interferon regulatory factor 2 (IRF2). METTL21C is a lysine methyltransferase expressed in skeletal muscle that activates NF-kB and may contribute to sarcopenia [33, 34]. ILRUN has been shown to inhibit interferon (IFN) regulatory factor 3 (IRF3)-mediated transcription, leading to decreased production of type-I IFN and tumor necrosis factor (TNF) cytokines [35]. In skeletal muscle, IRF2 can act as a negative regulator to suppress inflammation and improve cell viability. During the progression of OA progression, the expression of inflammatory genes increases in skeletal muscle. The ability for IRF2 to control the expression of genes involved in muscle inflammation suggests that it could be a target for new treatments for knee OA [36, 37].

Three genes associated with knee OA are involved in calcium transport and signaling: calcium voltage-gated channel subunit alpha1 E (CACNA1E), Mitochondrial Calcium Uptake 2 (MICU2), and Regulator of Calcineurin 1 (RCAN1). CACNA1E, a calcium channel regulatory gene, is essential for skeletal muscle contraction, development and regeneration. Human studies have found that the downregulation of CACNA1H, a related calcium channel gene, can induce skeletal muscle atrophy by disrupting calcium regulation. Aging alters the expression and activity of various calcium channels in skeletal muscle, which contributes to a reduction in muscle force generation and regeneration, this increased expression may be helping the working muscle [38]. MICU2 is a regulatory subunit of the mitochondrial calcium uniporter (MCU) complex, which controls the quantity of calcium enters the mitochondria. This mitochondrial calcium uptake is crucial for skeletal muscle health and performance. Its increased expression with knee OA suggests a need for more mitochondrial function with the increased knee OA severity [39]. RCAN1 inhibits calcineurin, an enzyme involved in regulating calcium signaling in the muscle related to mitochondrial function [40, 41].

Downregulated genes in knee OA belonged to several biologic pathways active in skeletal muscle. Sterile alpha motif domain containing 4A (SAMD4A) is an RNA-binding protein that regulates the mTORC1 signaling pathway, which is critical for muscle cell metabolism and potentially acts as a mediator between BCAAs and BDNF expression [42]. Long non-coding RNAs can influence muscle atrophy, but the specific function of Lnc-ZNHIT2-1, a long non-coding RNA, has not been established [43]. Expression levels for Lnc-ZNHIT2-1 were very low, resulting in a large amount of variability in our association estimate for this transcript. Dual Specificity Phosphatase 3 (DUSP3) is involved in regulating the MAPK/ERK pathway, which is important for cell proliferation and differentiation [44].

In addition to genes with well-understood biological functions related to knee OA, there are also genes with expression associated with knee OA, but their role in OA and their molecular function is not well understood. The most significantly associated differentially expressed gene, RPL29P11, is a pseudogene with unknown function. A pseudogene is a non-functional copy of an active gene (in this case, RPL29) that is not translated into proteins but can still be transcribed into RNA. The active gene, RPL29, encodes a ribosomal protein crucial for protein synthesis. Deletion of the active RPL29 gene in mice can inhibit angiogenesis (blood vessel formation), and this may be a part of the changes in muscle with knee OA [45, 46].

Our study has notable strengths and some limitations. Data collection for the SOMMA cohort was comprehensive, allowing for the adjustment for potential confounders of OA, including weight, waist circumference, and total abdominal adiposity. Since obesity and adipose tissue are strongly associated with OA, our adjustment for these confounders allows our results to be interpreted as largely independent of the well-known effect of obesity [24]. There are also some potential limitations. First, this is a community-based sample of older adult participants that could walk at a certain speed, and had no debilitating or life-threatening diseases, that would prevent them from participating in the SOMMA longitudinal epidemiologic study. Therefore, these results may not be generalized to a clinical population of knee OA subjects under medical care for knee OA. The muscle biopsy was obtained from the vastus lateralis muscle and the changes observed are limited to that muscle group and may not generalize to all muscles in the upper thigh. Also, there was one year from the time of the muscle biopsy until the knee radiographs, therefore changes may have occurred in the muscle due to the severity of the knee OA that was not captured in our study. Lastly, the extent to which our cross-sectional results reflect reverse causality (the influence of knee OA on muscle gene expression) cannot be ascertained; however, gene expression in muscle that responds to prevalent knee OA can help to identify biomarkers and potentially factors that influence OA progression after initial development.

In summary, knee OA in older adult participants is associated with expression of genes involved in pain sensitization, BCAA catabolism, the preservation of muscle function, calcium transport and storage, inflammation, and ECM remodeling. Additional longitudinal studies will determine how these genes affect the progression of the disease.

## Data Availability

All data are available online at sommaonline.ucsf.edu.

## Funding

SOMMA was funded by the National Institute on Aging (NIA) grant number R01AG059416. The knee osteoarthritis ancillary study was funded by NIA (R01AG070647, PI: Lane). Study infrastructure support was funded in part by NIA Claude D. Pepper Older American Independence Centers at University of Pittsburgh (P30AG024827) and Wake Forest University (P30AG021332) and the Clinical and Translational Science Institutes, funded by the National Center for Advancing Translational Science, at Wake Forest University (UL1 0TR001420).

## Supplemental Tables

Table S1. Participant characteristics stratified by sex.

Table S2. Differential gene expression significant (FDR ≤ 0.05) for either of the contrasts. Table S3. Sex-stratified differential gene expression for 16 genes associated in both contrasts.

## Notes

### Competing Interest Statement

The authors have declared no competing interest.

### Author Declarations

Source data were openly available to the public before the initiation of the study. Data can be obtained at sommaonline.ucsf.edu. This ancillary cross-sectional study enrolled SOMMA participants who returned for the first annual visit and consented to completing a standing knee radiograph. Participants provided written informed consent, and the study was approved by the Western Institutional Review Board Copernicus Group (no. 20180764) for all participating sites [8, 9]. Data from the November 2024 SOMMA release was analyzed.

